# Reduced Continuity Index with Proactive Esophageal Cooling Compared to Luminal Temperature Monitoring During Radiofrequency Ablation

**DOI:** 10.1101/2024.04.09.24305586

**Authors:** Catherine Lazarus, Jacob Sherman, Natalie Putzel, William Zagrodzky, Tiffany Sharkoski, Alex Ro, Jose Nazari, Westby Fisher, Erik Kulstad, Mark Metzl

**Affiliations:** Northwestern University, Evanston, IL, USA; Washington University in St. Louis, St. Louis, MO, USA; University of Southern California, Los Angeles, CA, USA; Colorado College, Colorado Springs, CO, 80946, USA; University of Pennsylvania, Philadelphia, PA, USA; NorthShore University Hospital, Evanston, IL, USA; University of Texas Southwestern Medical Center, Dallas, TX, USA

**Keywords:** Proactive esophageal cooling, Atrial fibrillation, Lesion continuity, Long-term freedom from atrial fibrillation, Pulmonary vein isolation, Radiofrequency ablation

## Abstract

**Background:** Proactive esophageal cooling is FDA cleared to reduce the likelihood of esophageal injury during radiofrequency ablation for treatment of atrial fibrillation (AF). Long-term follow-up data have also shown improved freedom from arrhythmia with proactive esophageal cooling compared to luminal esophageal temperature (LET) monitoring during pulmonary vein isolation (PVI). One hypothesized mechanism is improved lesion contiguity (as measured by the Continuity Index) with the use of cooling. We aimed to compare the Continuity Index of PVI cases using proactive esophageal cooling to those using LET monitoring.

**Methods:** Continuity Index was calculated for PVI cases at two different hospitals within the same health system using a slightly modified Continuity Index to facilitate both real-time calculation during observation of PVI cases and retrospective determination from recorded cases. The results were then compared between proactively cooled cases and those using LET monitoring.

**Results:** Continuity Indices for a total of 101 cases were obtained; 77 cases using proactive esophageal cooling and 24 cases using traditional LET monitoring. With proactive esophageal cooling, the average Continuity Index was 2.7 (1.3 on the left pulmonary vein, and 1.5 on the right pulmonary vein). With LET monitoring, the average Continuity Index was 27.3 (14.3 on the left, and 12.9 on the right), for a difference of 24.6 (p < 0.001).

**Conclusion:** Proactive esophageal cooling during PVI is associated with significantly improved lesion contiguity when compared to LET monitoring. This finding may offer a mechanism for the greater freedom from arrhythmia seen with proactive cooling in long-term follow-up.

## Introduction

Atrial fibrillation (AF) is a common form of cardiac arrhythmia increasingly treated with radiofrequency (RF) pulmonary vein isolation (PVI) ablation.^1^ As new technologies continue to emerge, improving procedural efficacy while maintaining procedural safety remains paramount. Procedural efficacy, as measured by long-term freedom from arrhythmia, has been shown to increase with improved contiguity of lesion placement.^2-4^ However, lesion contiguity is often limited by the need to pause and reposition the ablation catheter in response to temperature increases in the esophagus when using luminal esophageal temperature (LET) monitoring.

Temperature increases in the esophagus generally require cessation of RF delivery and result in delays in placing the adjacent lesion while awaiting return to equilibrium temperatures or performing repositioning of the catheter elsewhere in the left atrium.

An increasingly utilized alternative to LET monitoring is proactive esophageal cooling using a dedicated esophageal cooling device. Proactive esophageal cooling during RF ablation has been shown to reduce endoscopically identified esophageal lesions by 83% ^5^, and a recent analysis of 25,186 patients found a significant reduction in atrioesophageal fistulas (AEFs) with cooling.^6^ This approach is now also cleared by the US Food and Drug Administration (FDA) to reduce the likelihood of ablation-related esophageal injury resulting from RF cardiac ablation procedures.^7^ In addition to enhanced safety, recent data have shown improved long-term freedom from arrhythmia with proactive esophageal cooling when compared to LET monitoring during PVI. The long-term follow-up from the 120 patient IMPACT randomized controlled trial found a 3% (p=ns) absolute improvement, and a 513 patient review found a 14% absolute improvement (p=0.03) in freedom from arrhythmia at one year.^8,9^

A potential mechanism for an improvement in long-term freedom from AF with proactive esophageal cooling is via improvement in lesion contiguity, or lesion sequentiality.^2,3^ Lesion contiguity has been characterized using the Continuity Index, which is a quantification determined by the number of non-contiguous, or non-adjacent, lesions placed.^3^ Each non-contiguous lesion increments the Continuity Index, such that the higher the Continuity Index, the lower the overall contiguity of lesions. Cases with lower Continuity Indices have been shown to result in better isolation and greater freedom from arrhythmia when compared to procedures with higher Continuity Indices.^3^ Likewise, cases with higher Continuity Indices exhibit higher rates of gap reformation due to the rapid edema formation that occurs immediately following lesion placement, preventing subsequent lesions from achieving full thickness transmurality.^3^ Proactive esophageal cooling eliminates the need to pause RF lesion placement due to temperature rises in the esophagus, as no temperature monitoring is utilized with the system.

As such, proactive esophageal cooling may allow the Continuity Index to be lower than is otherwise possible when using LET monitoring. To test this hypothesis, we measured the Continuity Index of PVI cases and compared the results between cases using proactive esophageal cooling to cases using LET monitoring.

## Materials and Methods

### Study Design

This IRB reviewed and approved study included a prospective observational arm and a retrospective arm. Investigators recorded the Continuity Index prospectively in real-time during procedures performed by any of 4 operators performing ablations with proactive cooling. The same investigators then reviewed recordings of cases that had been completed prior to the health system adopting esophageal cooling, in which 3 of the same operators performed ablation with LET monitoring.

### Patients

Patients included those undergoing PVI procedures for the treatment of AF by any of four operators. AF types included paroxysmal, persistent, and long-standing persistent AF. Cases were selected via convenience sampling based on availability of investigators to observe the case. Otherwise, there were no exclusion criteria.

### Data Collection

In the prospective arm of the study, data were collected on cases employing proactive esophageal cooling in real time. The methods for calculating the Continuity Index in real time have been described previously ^10^, but in brief, investigators observed cases from the EP lab control room to monitor lesion placement sequence on the 3-D electroanatomical map during PVI procedures. Calculation of the Continuity Index was then performed as discussed below. To determine the Continuity Index of cases utilizing LET monitoring, recordings of cases completed prior to the adoption of proactive esophageal cooling were reviewed, and the Continuity Index for each case was calculated identically. Patient age, gender, and type of AF, was recorded.

### Procedure

RF PVI ablations were performed similarly to those of most procedures performed in the US, with patients under general anesthesia, and wide area circumferential PVI performed with the use of electroanatomical and three-dimensional geometry mapping using the Carto 3 mapping system and an irrigated ablation ST/SF™ catheter (Biosense Webster, Inc., Diamond Bar, CA, USA). Mitral isthmus lines, additional posterior wall isolation, and cavotricuspid isthmus lines were employed dependent on physician preference. A high power short duration technique was targeted, and low or no fluoroscopy approach was utilized. There were no other changes in the procedural approach besides the type of esophageal protection.

In the cases in which proactive esophageal cooling was employed, a dedicated cooling device (ensoETM^®^, Attune Medical, Chicago, IL) consisting of a closed-loop multi-lumen medical-grade silicone tube that circulates cold water at a temperature of 4°C was placed **(Figure 1)**. The device circulates distilled water at a rate of 2.4 L/minute between the device and an external heat exchanger. The device replaces the need for LET monitoring and is positioned similarly to an orogastric tube, avoiding interference with procedural workflow. Device placement is confirmed by either fluoroscopy or intracardiac echocardiography.

**Figure 1:**
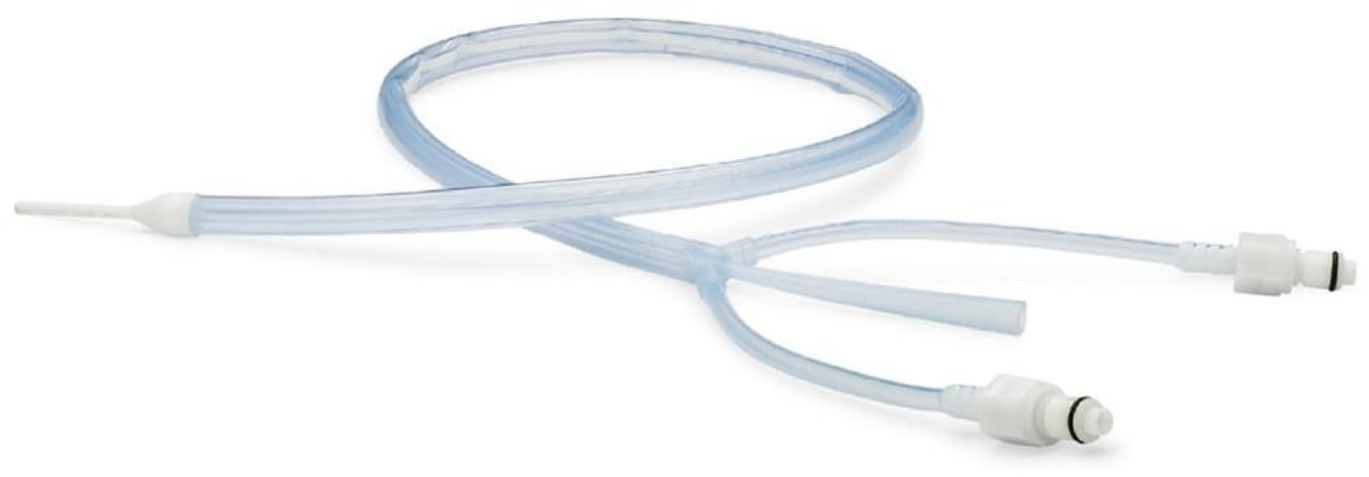
Proactive esophageal cooling device.

In the cases using LET monitoring, a multi-sensor probe (Circa S-Cath; Circa Scientific, Inc., Englewood, CO, USA) was employed. Energy delivery was discontinued when the maximum LET on any sensor of the multi-sensor probe rose by more than 0.2 °C/s or exceeded 39 °C.

### Continuity Index Calculation

The original description of the Continuity Index divided the left atrium into segments, and incremented the Index by the number of segments the catheter was moved to place a noncontiguous lesion **(Figure 2)**.^3^

**Figure 2:**
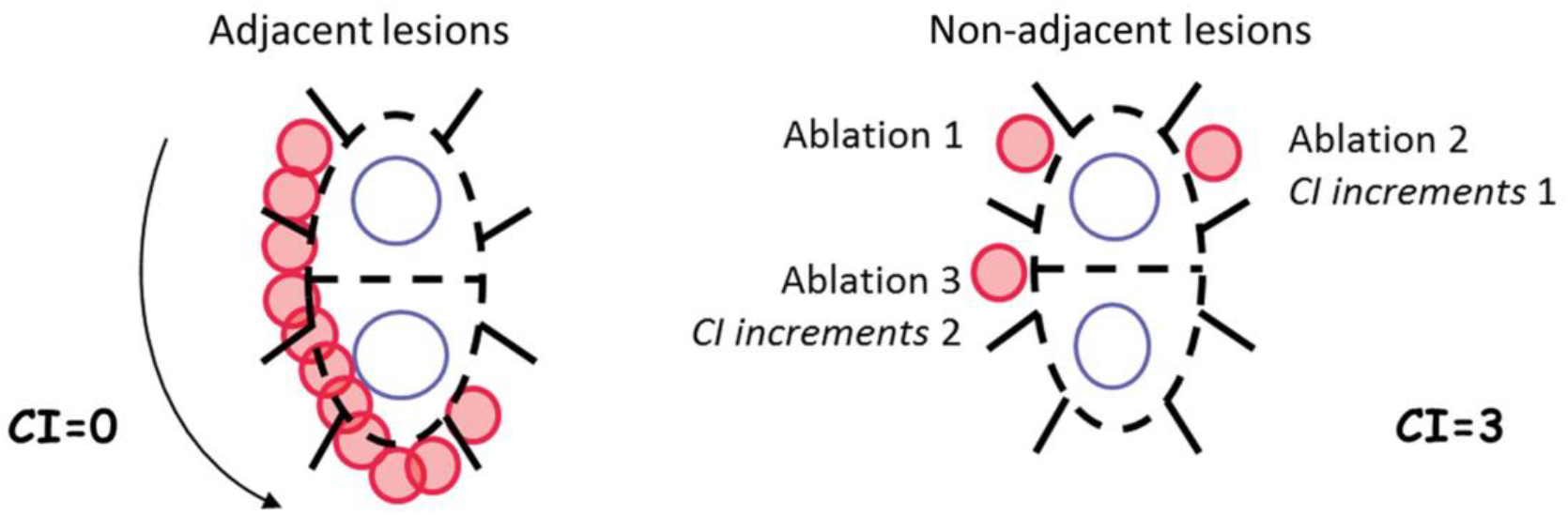
Original Continuity Index.^3^ (With permission.)

In order to better facilitate standardization and improve ease of use in real-time, we utilized a slight modification, incrementing the Index by one unit for each lesion that does not border a previous lesion, regardless of distance placed from the previous lesion **(Figure 3)**. This modification avoided the ambiguity inherent with estimating exact demarcations of segments of the left atrium, which otherwise would add variability and increase subjectivity in determination of the Continuity Index. This modified approach was followed to calculate the Indices for both the prospective and retrospective groups. As an example, if an operator deployed every lesion adjacent to the previous lesion without any discontinuity, the final Continuity Index would be 0. Each lesion placed non-contiguously would increment the Index by one unit.

**Figure 3:**
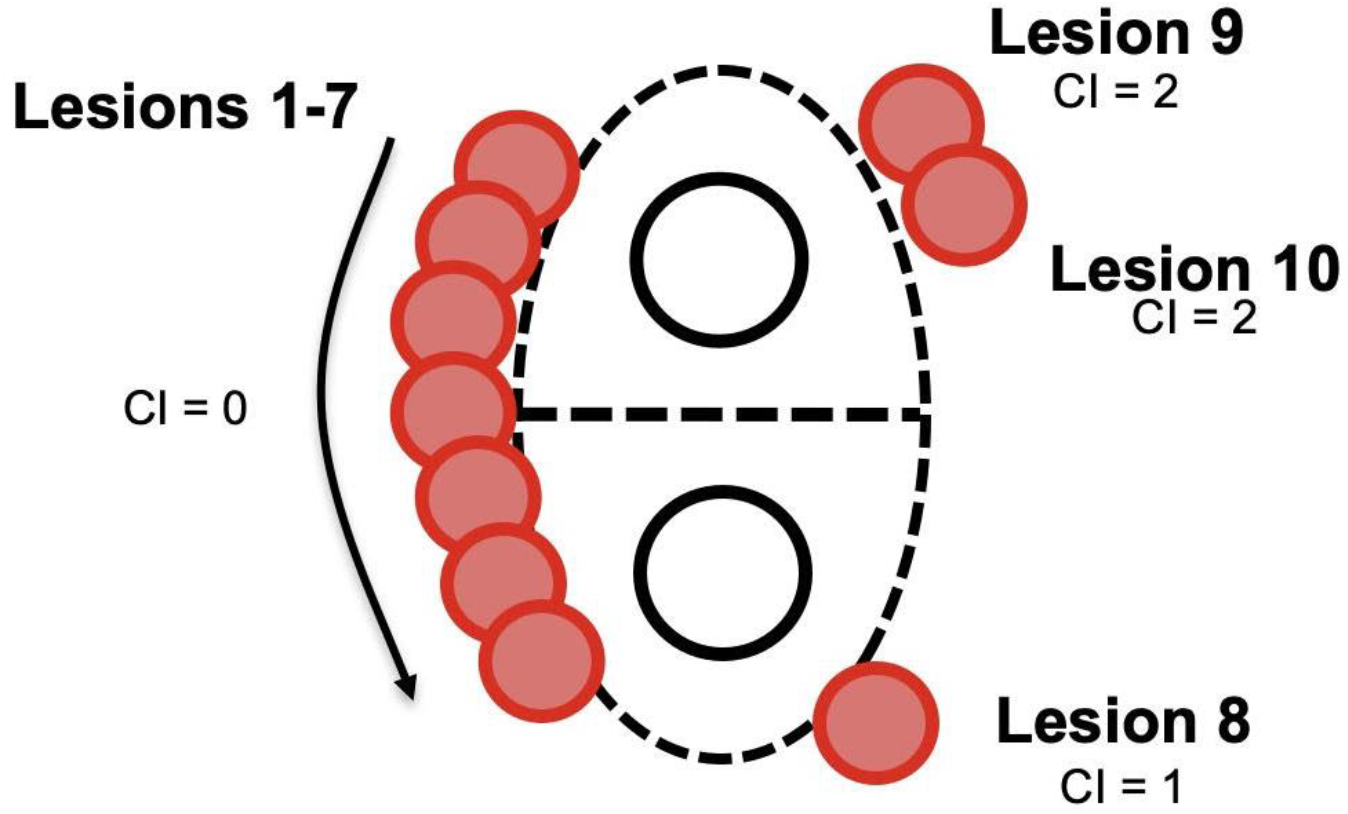
Minor modification to the Continuity Index (CI). “Lesions 1-7” represent a Continuity Index of 0, as each lesion is directly adjacent or overlapping with the previous lesion. The deployment of “Lesion 8” makes the Continuity Index 1, as there is a discontinuity between lesions 7 and 8 with no direct overlap. Similarly, “Lesion 9” further increased the Continuity Index to 2 because it is not contiguous with “Lesion 8.” There is no change with the addition of “Lesion 10,” as it is directly adjacent to “Lesion 9.”

### Data Analysis

The mean Continuity Index was calculated for each group, and results compared between the LET group and the esophageal cooling group. Data were analyzed using SPSS (IBM SPSS Statistics for Windows, Version 29.0. Armonk, NY: IBM Corp.) Continuous variables are presented as mean±SD.

## Results

### Patient Demographics

The Continuity Index was determined for a total of 101 cases. Of these, 77 cases using proactive esophageal cooling were obtained in real-time during cases performed from June 2023 to December 2023. Another 24 cases were determined via review of recorded cases performed using LET monitoring prior to adoption of proactive esophageal cooling (which occurred in January 2019). Patient demographics for both treatment groups were similar. The average age for the proactively cooled group was 69.9 (SD ± 9.0) years while the average age was 69.2 (SD ± 10.0) years for the LET monitored group. Likewise, there was no significant difference in gender between the two groups, with the proactively cooled group consisting of 55.8% male and the LET monitored group consisting of 62.5% male.

### Continuity Index

Cases that used proactive esophageal cooling had a mean Continuity Index of 1.5 (SD ± 2.3) for the right pulmonary veins and 1.3 (SD ± 1.7) for the left pulmonary veins, for a mean total Continuity Index of 2.7 (SD ± 3.7)**(Table 1)**. Cases using LET monitoring had a mean Continuity Index of 12.9 (SD ± 3.9) for the right pulmonary veins and 14.3 (SD ± 5.1) for the left pulmonary veins, for a mean total Continuity Index of 27.3 (SD ± 8.2), representing a significant order of magnitude difference (p < 0.001) **(Figure 4)**. A total of 4 patients (5%) in the esophageal cooling group had a Continuity Index above 6 on either side, compared to all patients (100%) in the LET monitoring group.

**Table 1:** Detailed Continuity Index parameters.

|  | Continuity Index |  |  |
| --- | --- | --- | --- |
|  | Cooling<br>(n=77) | LET<br>monitoring<br>(n=24) |  |
| **Left side** |  |  |  |
| mean | 1.7 | 14.3 | $p = <0.001$ |
| SD | 1.7 | 5.1 |  |
| min | 0 | 7 |  |
| max | 6 | 28 |  |
| **Right side** |  |  |  |
| mean | 1.5 | 12.9 | p =<0.001 |
| SD | 2.3 | 3.9 |  |
| min | 0 | 6 |  |
| max | 9 | 22 |  |
| **Total** |  |  |  |
| mean | 2.7 | 27.3 | p =<0.001 |
| SD | 3.7 | 8.2 |  |
| min | 0 | 15 |  |
| max | 14 | 50 |  |

**Figure 4:**
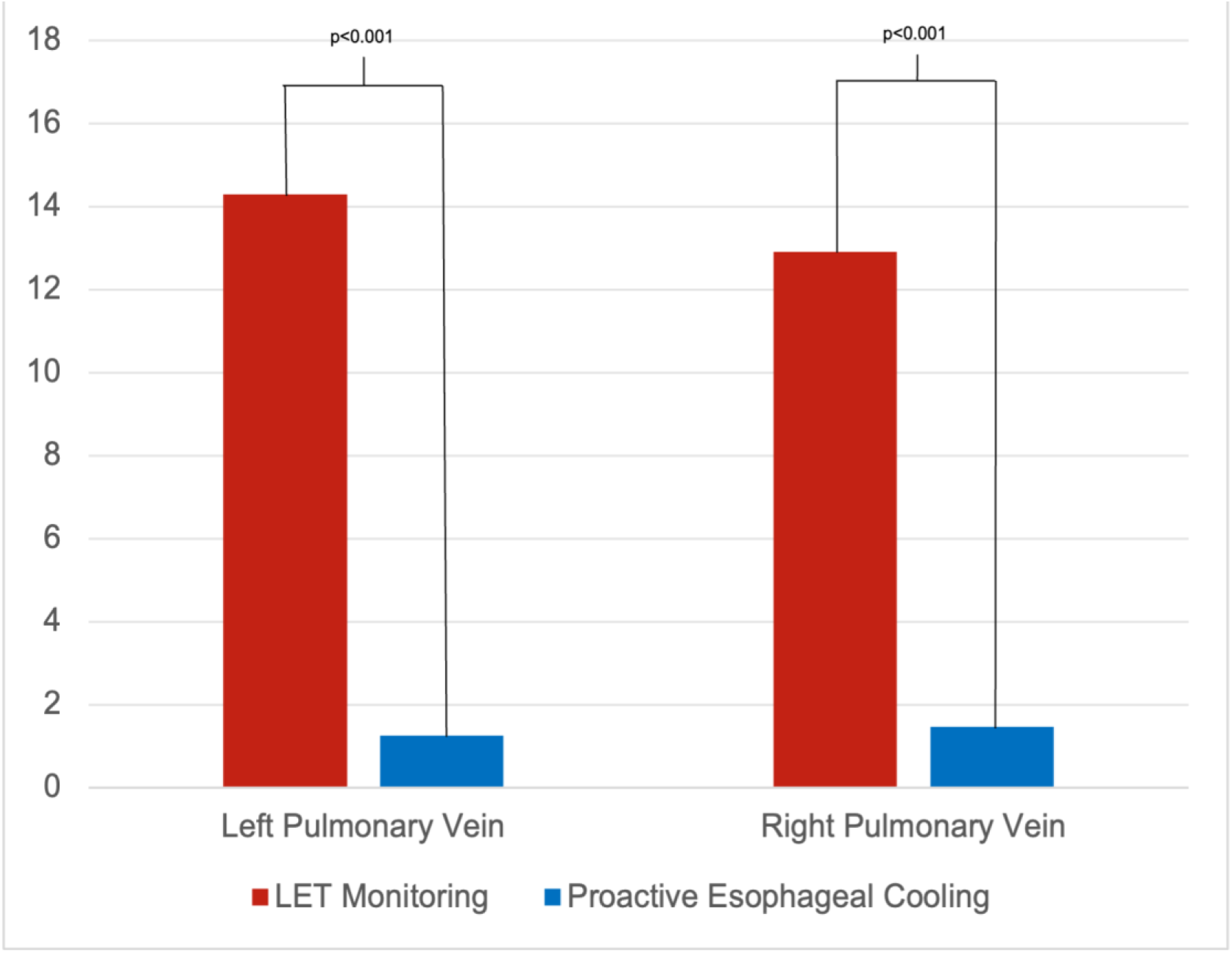
Comparison of Continuity Index (modified) between LET monitoring and proactive esophageal cooling.

## Discussion

This is the first study to compare the Continuity Index obtained with different methods of esophageal protection, finding that the Continuity Index is significantly lower (by roughly an order of magnitude) when using proactive esophageal cooling than with LET monitoring. Cases utilizing proactive esophageal cooling had a mean Continuity Index of 2.7, whereas those utilizing LET monitoring had a mean Continuity Index of 27.3 (p < 0.001). Proactive esophageal cooling enabled significant improvement in lesion contiguity by eliminating interruptions of RF delivery that would otherwise be encountered in cases employing LET monitoring.

The formal measure of contiguity via a Continuity Index was proposed by Kautzner et al. in the EFFICAS-II study in 2015.^3^ In this study, Continuity Indices above 6 (signifying a lower lesion contiguity) corresponded to a significantly diminished likelihood of preserving long-term freedom from AF when compared to cases with Continuity Indices below 6 (signifying a higher degree of lesion contiguity).^3^ Specifically, PV lines isolated initially with a Continuity Index below 6 had a 98% (56/57) chance of remaining isolated, as compared with PV lines with a Continuity Index ≥ 6, which had only a 62% (21/34) chance of remaining isolated (p < 0.001). Further methods of quantifying lesion sequentiality have since been described by Jankelson et al., where arrhythmia-free survival was significantly higher in patients with greater lesion sequentiality (96% vs 76%, p = .01).^2^ The higher occurrence of gaps is due to at least two factors. The first is the partial transmurality that results from premature cessation of RF energy after detecting an intra-esophageal temperature rise. The second is the fact that edema begins to form around each lesion soon after lesion placement.^11-13^ This growing edema hinders subsequent transmural lesion formation at adjacent positions, because once the operator returns to the site to place an adjacent lesion, the tissue is no longer the same thickness and will require different energy parameters to achieve transmurality.^4^ In cases with LET monitoring, it is common practice to reposition the ablation catheter to other regions of the atrium to prevent heat stacking and overheating.^14^

The temporal aspect of lesion placement is important, since lesions placed immediately are more likely to exhibit transmurality-associated unipolar electrograms as evidence of lesion transmurality.^4^ In cases with lower Continuity Indices, there is less time for edema to form around lesions providing a higher likelihood of achieving transmurality due to more consistent tissue thickness.^3^ In proactively cooled cases, there is no compulsion to interrupt RF delivery, as the esophagus is proactively cooled to 4° C and no temperature monitoring is required. This approach additionally offers greater safety, since in contrast to LET monitoring, which has not demonstrated benefit in reducing esophageal injury ^15-17^, proactive esophageal cooling has demonstrated significant reduction in both esophageal injury and AEF rates.^5,6^ As such, LET monitoring remains unapproved by the FDA, whereas proactive esophageal cooling is FDA cleared to reduce the likelihood of ablation-related esophageal injury resulting from RF cardiac ablation procedures.^7^

### Study Limitations

Given the nature of this study, investigators were not blinded to the mode of esophageal protection utilized. However, objective endpoints of lesion placement order are unlikely to be influenced by knowledge of mode of esophageal protection. Cases with LET monitoring were performed by 3 of the 4 operators using proactive esophageal cooling, but this study otherwise followed the same operators with consistent procedural approaches, operating within the same hospital system, and serving the same patient demographic over the time period. Long-term follow-up comparison of procedural outcome is not available for the cases reviewed in this analysis; however, abundant data, including prospective randomized controlled trial data ^8^, as well as larger retrospective data from this same site ^9^, support the improved efficacy from proactive esophageal cooling, and ongoing randomized trials (NCT04577859) will continue to quantify this effect size.

## Conclusion

Proactive esophageal cooling during PVI is associated with significantly improved lesion contiguity when compared to LET monitoring. This finding may offer a mechanism for the greater freedom from arrhythmia seen with proactive cooling in long-term follow-up.

## Data Availability

Data available from corresponding author upon request.

